# Mapping polygenic risk scores to brain cells: insights for neurodegenerative diseases

**DOI:** 10.64898/2026.10.08.26365054

**Authors:** Emily Simmonds, Anna McKeever, Ümran Yaman, Ganna Leonenko, Kerbez Kimatova, Philip Taylor, Siddharthan Chandran, Mina Ryten, Baljit Khakh, John T O’Brien, Tara Spires-Jones, Dervis A. Salih, Valentina Escott-Price

**Author notes:** Correspondence to: Valentina Escott-Price, Full address Hadyn Ellis Building, Maindy Road, Cardiff, United Kingdom, CF24 4HQ.

## Abstract

Neurodegenerative diseases (NDDs) are driven by multiple related yet distinct pathological processes that evolve over decades, resulting in substantial clinical and neuropathological heterogeneity. Postmortem examinations of older individuals frequently reveal multiple coexisting neuropathologies, however, the underlying pathological processes cannot be easily assessed directly in living individuals. To quantify pathology-specific genetic signatures, we analysed a spectrum of NDDs using polygenic risk scores (PRSs) in the UK Biobank, restricting analyses to genes reported to be expressed in major brain cell-types, including microglia, astrocytes, oligodendrocytes, and neurons. We found substantial overlap among purportedly cell-type specific gene sets (21-29%), with *APOE* or neighbouring genes from the *APOE* locus represented in all gene sets, indicating that current cell-type specific annotations capture considerable shared genetic variation across cell-types. Jointly, cell-type specific PRSs explained a large proportion (60-93%) of the genetic liability for NDDs in which dementia is a major clinical manifestation. Alzheimer’s disease (AD) showed associations across all cell-types, with oligodendrocyte- and astrocyte-based PRSs demonstrating strong effects independent of *APOE*. AD PRS and *APOE* status independently differentiated Parkinson’s disease (PD) with dementia from PD without dementia. PD without dementia and amyotrophic lateral sclerosis (ALS) exhibited distinct genetic profiles, with limited overlap with other NDDs. Collectively, these findings reveal both shared and disease-specific genetic contributions across neurodegenerative disorders and demonstrate that currently available cell-type specific gene sets capture substantial overlapping genetic risk. The imperfect alignment between clinical phenotypes and underlying pathology, coupled with the extensive overlap of gene expression across these cell types, underscores the persistent challenges in resolving the genetic architecture of neurodegenerative diseases and elucidating their underlying biological mechanisms.

## Introduction

The critical need for earlier and more accurate diagnosis of neurodegenerative diseases has driven the integration of genetic approaches, primarily through polygenic risk score analyses, with plasma and cerebrospinal fluid biomarkers. However, challenges such as mixed pathologies, especially in aged individuals, and frequent misdiagnosis are insufficiently addressed, and are important considerations for the translation of polygenic risk scores to clinical practice.

Neurodegenerative diseases often present with overlapping symptoms and underlying pathological changes which can make accurate diagnosis complicated. Misdiagnosis is a persistent challenge, with studies indicating that a considerable number of patients initially diagnosed with one NDD are later reclassified upon neuropathological examination or disease progression ^1^. The current accuracy of the clinical diagnosis of Alzheimer’s disease (AD) has sensitivity of the order of 70-90% ^2^, but with lower (50-70%) specificity, though this accuracy can be increased when utilising plasma biomarkers for diagnosis ^3^. However, diagnostic uncertainty is compounded by the frequent co-occurrence of multiple neurodegenerative pathologies within the same individual ^4^. For instance, mixed pathologies involving AD and dementia with Lewy bodies (DLB) are common in aged adults ^5^ and may influence clinical presentation and treatment response ^6–8^.

Genome-wide association studies (GWAS) have identified numerous common genetic variants associated with increased disease risk; however, each variant individually confers only a modest effect. To address this, polygenic risk scores (PRS) have emerged as a useful tool to quantify an individual’s cumulative genetic liability by aggregating the effects of multiple risk alleles across the genome ^9^. In the context of neurodegeneration, PRS have shown value in enhancing risk prediction ^10–12^, elucidating shared genetic architecture across diseases, and identifying individuals at elevated risk prior to clinical onset. Recent studies highlight the potential of polygenic risk to capture shared molecular signatures, such as inflammatory pathways, that transcend traditional diagnostic boundaries and may potentially enable the distinction between sub-groups within diagnostic groups ^13^. Despite their potential, the clinical utility of PRS remains unrealized owing to several challenges. First, the predictive accuracy of PRS is contingent upon the size and ancestral diversity of the GWAS datasets from which they are derived. Secondly, since common variant genetics explains only a portion of SNP-based heritability, the prediction accuracy by PRS is, and will remain, limited ^14^. Finally, the imprecision of clinical diagnosis carried through GWAS, due to its design, further reduces the predictive accuracy of PRS.

Microglia, immune cells in the central nervous system (CNS), play a vital role in maintaining neural homeostasis and responding to injury or disease. In neurodegenerative diseases microglia undergo changes in state that can either mitigate or exacerbate disease progression. Emerging evidence suggests that microglial activation is not a uniform response but rather a spectrum of context-dependent states, ranging from beneficial to harmful phenotypes ^15,16^. A microglia gene-set has been shown to explain 69-84% of SNP-based heritability using only 3% of SNPs in AD ^17^. Other cell-types have also been shown to be relevant to neurodegenerative disorders ^18^. Synaptic dysfunction is a central and early feature of many neurodegenerative diseases. Synapses are critical for neuronal communication, plasticity, and network integrity, and their loss strongly correlates with cognitive decline ^19^. Moreover, recent studies have highlighted the role of immune-mediated synapse loss in accelerating synaptic dysfunction in early disease stages ^19^. Astrocytes are the most abundant glial cells in the central nervous system and are essential for maintaining neuronal health through regulation of synaptic transmission, metabolic support, and blood-brain barrier integrity ^20^. Astrocytes are also implicated in synapse formation and removal and in crosstalk with microglia in mediating neuroinflammation and neurodegeneration ^21,22^. Oligodendrocytes are also abundant glial cells that generate myelin to insulate axons for rapid nerve conduction, and provide critical metabolic support to neurons, supporting adaptive neural plasticity ^23^. Oligodendrocytes show functional changes in AD ^24^, contribute to the development of pathology ^25,26^, and are primarily affected in neurodegenerative diseases such as multiple sclerosis (MS) and multiple system atrophy (MSA). These findings underscore the importance of microglia, synapse, astrocyte and oligodendrocyte involvement in neurodegeneration and highlight their potential as therapeutic targets for modulating disease trajectories. Here we examine cell-type specific polygenic risk across multiple neurodegenerative diseases (NDDs) in a large, clinically diagnosed cohort focusing on four major cell types affected in NDD: microglia, astrocytes, oligodendrocytes and neurons with a focus on neuronal genes involved in synaptic function.

A recent study has shown that cell-type specific AD PRS are associated with distinct disease processes in AD uncovering the impact of cell-type specific genetic risk on AD endophenotypes in an autopsy dataset ^27^. We expand upon this work by investigating the cell-type specific effect in living individuals and across all NDDs, not just AD.

This study aims to investigate the strength of association and report prediction accuracies between NDDs and PRS defined by the whole genome and cell-type specific genes expressed in microglia, synapse, astrocytes and oligodendrocytes. The association between cell-type specific PRS and each NDD will be modelled jointly, to account for overlap between gene-sets, and separately, to give an overall measure of association for each cell-type irrespective of other cell-types. To investigate whether disease-specific and/or cell-type specific PRS have an advantage of improved disease prediction accuracy or between disease specificity, we test whether NDD phenotypes can be predicted by PRS generated with SNP effect sizes from other NDDs. Finally, we also assess whether any cell-type specific PRS are associated with neurodegenerative biomarkers (neurofilament light (NfL) and glial fibrillary acidic protein (GFAP)). We used the UK Biobank dataset ^28^, which includes extensive genotyping and longitudinal health records, enabling our study to explore NDDs disease risk in a variety of PRS settings.

## Materials and methods

### Data Description

UK Biobank (UKB) ^28^ is a large prospective cohort of individuals from the UK recruited between 2006 and 2010. UKB access was under application number 109607 and data were dispensed in Oct 2024. UKB contains extensive phenotype and imputed genetic data required for this analysis.

UKB has derived diagnostic phenotypes, in particular for dementia (N=8,294), Alzheimer’s Disease (AD) (N=3,729), Parkinson’s disease (PD) (N=3,681), frontotemporal dementia (FTD) (N=261), vascular dementia (N=1,805) and Amyotrophic Lateral Sclerosis (ALS, sometimes referred to as Motor Neuron Disease) (N=631), codes 42018, 42020, 42032, 42024, 42022 and 42028 respectively which contain algorithmic combinations of baseline assessment data, self-reported outcomes, hospital data and death register records. Currently there is no specific code for Dementia with Lewy Bodies (DLB) in UK Biobank. The dementia phenotype captures individuals with “unspecified” dementia who are not captured by other NDD phenotypes. Diagnosis is measured at multiple time points; here we take the first recorded diagnosis. Controls are defined as any individuals in UK Biobank who were not diagnosed with any of the neurodegenerative diseases above and multiple sclerosis (MS) (N=395,173). The percentage of individuals with multiple diagnoses was noted. We included individuals recorded as Caucasian based on the field 22006 in the analysis. We also extracted covariates such as age, sex and genetic principal components.

### Discovery summary statistics for Polygenic Risk Scores

We used the latest and the largest publicly available European GWAS summary statistics without UK Biobank for AD, PD, DLB, FTD, ALS and ageing/longevity. For the AD polygenic risk score, Kunkle et al. ^29^ the largest GWAS with clinical phenotyping (N cases=21,982 and N controls=41,944) was used and a separate score was computed excluding the *APOE* region (chr19: 44.4-46.5Mb). For PD the Nalls et al. ^30^ summary statistics were used (N cases=37,700 and N controls=1.4million), excluding individuals in the UKB. Summary statistics for FTD (N cases=3,348 and N controls=9,390), DLB (N cases=2,591, N controls=4,027) and ALS (N cases=27,205, N controls=110,881) are taken from Mishra et al. ^31^, Chia et al. ^32^ and van Rheenen et al. ^33^ respectively. Ageing/longevity GWAS includes individuals surviving to the 90^th^ percentile of life and controls whose age at death corresponds to the 60^th^ percentile (N cases=11,262, N controls=25,483) ^34^. Due to the comorbidity of vascular dementia with cardiovascular disease, we also computed a coronary artery disease (CAD) PRS using CAD GWAS (Ncases=60,801, Ncontrols=123,504) taken from Nikpay et al. ^35^. This CAD PRS is used as a proxy for vascular dementia as it has been shown to be associated with vascular dementia ^36^, and we were unable to use a vascular dementia GWAS as it includes UK Biobank individuals. We only used CAD-PRS to inform vascular dementia.

Polygenic risk score-continuous shrinkage (PRS-CS) ^37^ was used to estimate posterior SNP effect sizes under linkage disequilibrium (LD) modelling. We employed PRS-CS with default settings, and the global shrinkage (phi) parameter was optimised based on the data. We estimated LD between SNPs using the UKB European reference panel made available on the PRS-CS GitHub. These adjusted weights were then used to weight the number of risk alleles an individual has in the UKB data using the “--score” command in Plink.v2 ^38^. UKB data imputed with the Haplotype Reference Consortium reference panel by UKB were quality controlled by removing SNPs imputed with INFO (imputation accuracy) score < 0.4, minor allele frequency (MAF) < 1%, SNP missingness > 5%, individual missingness > 5% and SNPs not in Hardy-Weinberg Equilibrium (p < 1e-6). The remaining SNPs were included into the genetic risk score. The PRS was adjusted for 15 principal components (by regressing off the PCs) and standardised.

This method was applied for all PRS in this analysis, to ensure that the PRS calculation approach was consistent for all NDDs and across all cell-type specific gene sets. We did not use the UK Biobank PRS release ^39^, because they do not include PRS for all NDDs investigated, specific cell-types or all individuals.To focus on specific biological functions, we have utilised several defined biological gene-sets relevant to neurodegeneration. Our choice of gene sets was guided by the largest and most cited sources where possible, as advised by the specialists in the field. Gosselin et al. ^40^ defined microglia regions based on GWAS signatures and epigenetic/gene regulatory data. The genes within these microglia regions were extracted, SNPs within the start and end position of these genes plus a window of 35kb upstream and 10kb downstream were identified, and the polygenic risk score was restricted to these specific SNPs to create a microglia specific PRS. A synapse ^41^ gene-set has been defined using Gene Ontology (GO) terms for synaptic locations and processes, in this case SNPs from 35kb upstream and 10kb downstream of the gene were included. An astrocyte specific gene-set was defined by selecting genes within the top 10% of expression specificity by ranking genes based on differential expression for astrocytes ^27,42^, a window of 35kb upstream and 10kb downstream around the gene start and end positions was again used. Note that the resulting astrocyte gene-set did not include *APOE*. A second astrocyte gene-set found in mice ^43^ by utilising gene expression and astrocyte morphology data and comparing these genes with astrocytic differentially expressed genes from AD mouse models, was also assessed and compared which did contain *APOE*. An oligodendrocyte gene-set list was based on annotations in ROSMAP data by Mathys et al. 2023 ^44^ and Xiong et al. 2023 ^45^. Genes preferentially expressed by oligodendrocytes in controls were used from the ROSMAP single-nucleus expression data, where genes with p-adj < 0.05 in oligodendrocytes compared to other cell types were used. In addition, we looked at genes preferentially expressed by oligodendrocytes in late stage AD and tested the resulting PRS for association with AD only. The PRS for microglia, synapse, astrocyte and oligodendrocyte gene-sets were computed based on AD, PD, DLB, FTD, ALS and ageing/longevity GWAS summary statistics. In addition, a “no gene-set” PRS was introduced, which is calculated for SNPs from the rest of the genome excluding microglia, synapse, astrocyte and oligodendrocyte gene-sets and the *APOE* region (chr19:44.4-46.5Mb, hg19).

As some genes may have important roles in multiple cell types, we allowed the overlap of cell-type specific genes. This was necessary as the cell-type gene-sets were defined independently by separate research groups, without consideration of other cell-types, therefore there are genes which are assigned to multiple gene-sets. To ensure the gene-sets are as comparable as possible, we exclude *APOE* from all gene-sets (chr19:44.4-46.5Mb, hg19). We accounted for the *APOE* effect by including two variables representing the numbers of *APOE*-e4 and *APOE*-e2 alleles in the regression model. Figure 1 shows all phenotypes and PRS tested in this study.

**Figure 1.**
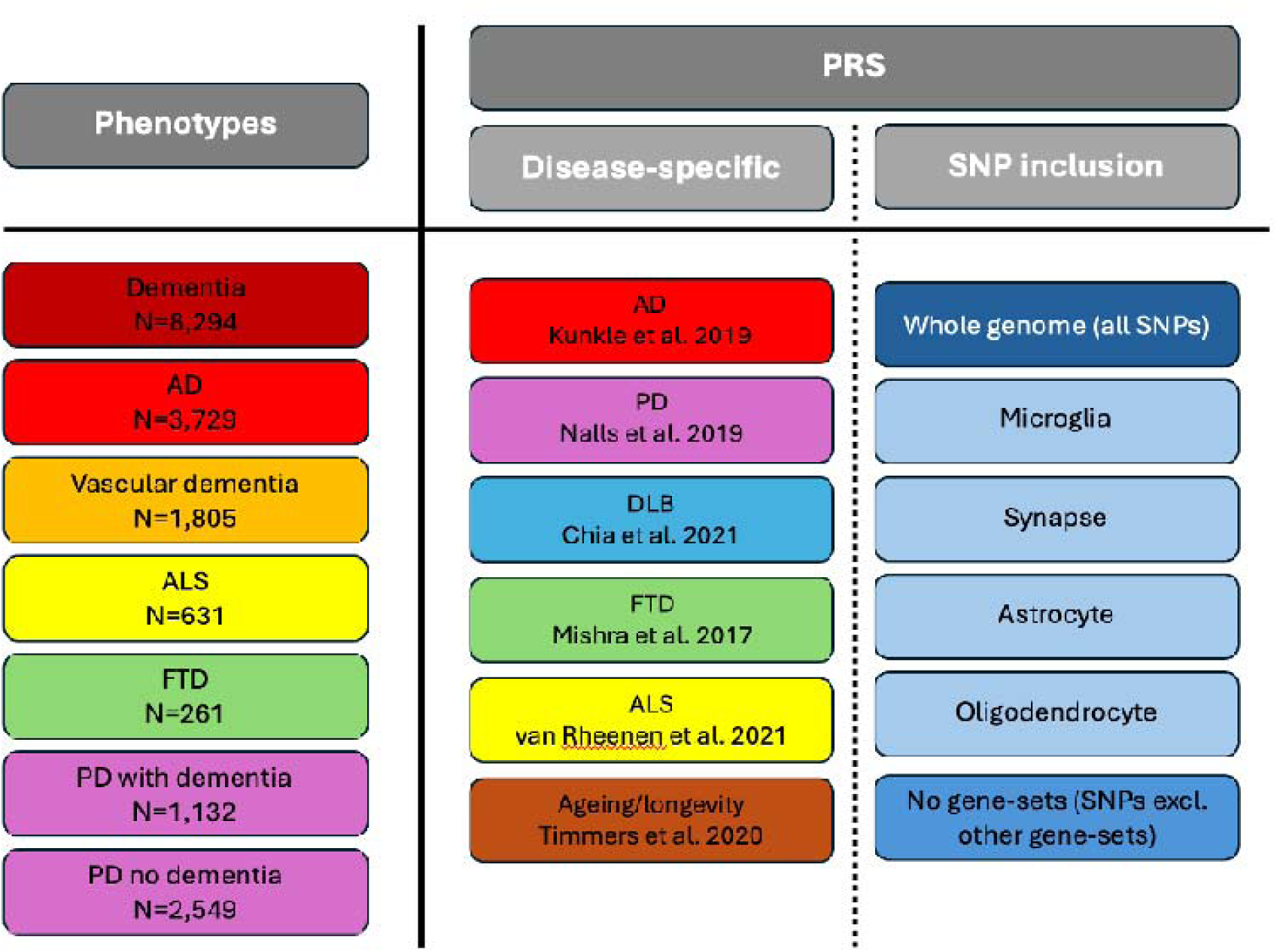
Study Design showing all Phenotypes and polygenic risk scores (PRS) in this study. PRS=polygenic risk score, SNP=single nucleotide polymorphisms, AD= Alzheimer’s Disease, ALS=amyotrophic lateral sclerosis, FTD=frontotemporal dementia, PD=Parkinson’s Disease, DLB=dementia with Lewy bodies. The colours for disease boxes highlight a specific disease, for SNP inclusion boxes, the dark blue is for all SNPs, the pale blue is for cell-type specific SNPs and the mid blue is for the gene-set containing all SNPs not in the four gene-sets.

### Statistical Analysis

We employed Lasso regression ^46^ to determine which combination of cell-type PRS best explain each phenotype and cross-validated the results splitting the data into a training and test set with an 80/20 split. Lasso regression enables the correlation between gene-sets to be accounted for, and determines the association over and above *APOE*. We further used Cox regression for each NDD-PRS pair separately to investigate the strength of association between the each cell-type PRS and phenotype, with the time variable coded as either age at onset (first recorded diagnosis), age in 2025 or age at death. The Cox model accounts for age which is important, since disease stage may differ by age and therefore, be influenced by different cell-types. Beta (log(HR)), R2 and area under the receiver operating characteristic curve (AUC) are reported for each model, see Supplementary Methods for details. Logistic regression is used to compare association of PRS between different phenotypes, i.e. PD with dementia vs PD without dementia, to be consistent with Cox models, Beta is reported to demonstrate the strength and direction of association. Reported p-values of PRS association with a disease are corrected for multiple testing using an FDR correction. All models were adjusted for genetic sex, and only individuals of Caucasian ancestry are included.

The proportion of variability captured by the cell-type specific PRS was identified as *R^2^_cell-type_/R^2^_whole genome_*, where the numerator is calculated from the logistic regression model with a PRS including SNPs in all four cell-types as a predictor and the denominator is calculated for the whole genome PRS (all including *APOE* as per gene-set definition) as a predictor.

Finally, we assessed the association between commonly used neurodegenerative biomarkers measured for Biobank individuals, namely GFAP and NfL, with each of the cell-type specific PRS using linear regression. For these analyses, Beta coefficients are reported as measures of the strength and direction of association.

## Results

### Dementia Diagnosis Co-occurrence in UKB

Table 1 shows the number of individuals in each of the neurodegenerative disease groups (based on hospital inpatient records, baseline assessment, self-report and death register data) and the control group. It also presents the number of individuals with only this diagnosis, the number of males and females, the *APOE*-e4 frequency, the mean AAO in each group, the number of people within these groups who have died and the mean age at death. The age in controls is the age in 2025 or age at death. The mean age at diagnosis is notably lower in ALS, FTD and PD, and the control age is comparable to the neurodegenerative groups. The *APOE*-e4 frequency is highest in AD and lowest in ALS and controls.

**Table 1.**
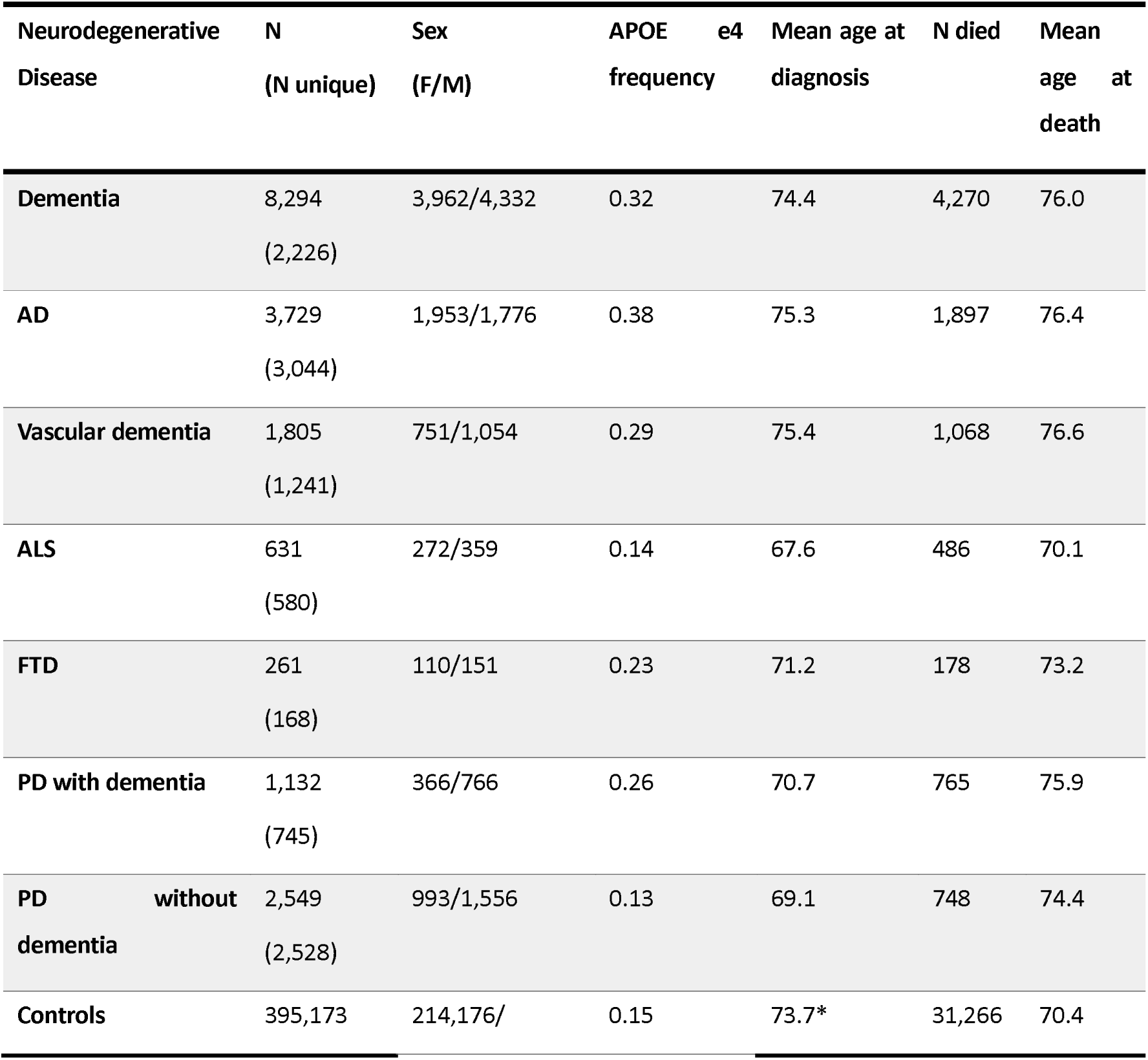
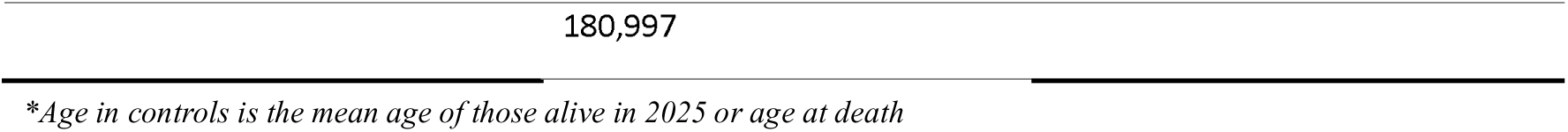
Number of Individuals within each neurodegenerative disease and control group.

Supplementary Table 1 shows the number of co-occurring dementia diagnoses (based on hospital inpatient records, baseline assessment, self-report and death register data) in UKB, with the percentage of total diagnoses. There is a high co-occurrence between PD with dementia and AD diagnoses (22.2%), vascular dementia and AD (23.6%), and 17.6% of individuals diagnosed with FTD are also diagnosed with AD. There is no overlap between PD without dementia and other neurodegenerative diseases, except for a small overlap with ALS, notably both these diseases do not have dementia as a symptom.

### Cell-type gene-set overlap

Table 2 presents the number of genes and SNPs in each gene-set, excluding the *APOE* region, as well as the percentage of SNPs out of all available SNPs. Supplementary Figure 1 is a Venn diagram of the SNP overlap between the cell-type specific gene-sets (see Supplementary Figure 2 for the gene overlap), all gene-sets in the Venn diagrams contain *APOE* as per the gene-set definition. Clearly, there is substantial overlap between gene-sets, this is due to research groups defining cell-type specific gene-sets independently. They are not specified considering any other cell-types, and several genes are expected to have diverse functions and therefore may be relevant to multiple cell-types.

**Table 2.** Number of genes and SNPs in each cell-type specific gene-set.

| Gene-set | Number of Genes | Number of SNPs (% compared to all SNPs) |
| --- | --- | --- |
| Microglia | 758 | 135,974 (2.3%) |
| Synapse | 1,061 | 366,707 (6.2%) |
| Astrocytes | 755 | 168,032 (2.8%) |
| Oligodendrocytes | 1,743 | 396,107 (6.7%) |

*Note that all gene-sets exclude APOE region (chr19:44.4-46.5Mb, hg19)*

Note that the proportion of SNPs are based on SNPs in UK Biobank, however, not all these SNPs are in the PRS as they must also be present in the summary statistics and in the reference data used to estimate linkage disequilibrium (LD) for PRS-CS. The proportion of SNPs in each gene-set is consistent across all summary statistics. We used gProfiler2 to run a gene enrichment analysis for each of the cell-type specific gene-sets, to show the function of the genes included in these gene-sets, the top 20 terms for each gene-set are displayed in Supplementary Figure 3. Despite the apparent SNP overlap between gene-sets, there is little overlap in the top 20 gene functions for each gene-set, suggesting that the overall gene-sets are capturing different processes.

When comparing astrocyte-enriched gene-sets from Endo et al. ^43^ and Mathys et al. ^42^, the prediction accuracy is similar across all NDDs (see Supplementary Table 2). The Endo et al. gene-set contains the *APOE* gene with 190 SNPs in the *APOE* region and Mathys et al. contains no SNPs in the *APOE* locus. As *APOE* is known to be expressed in astrocytes in individuals without AD ^47^, the Endo et al. gene-set is taken forward in this paper with *APOE* modelled independently of the PRS.

Since the cell-type specific gene-sets show substantial overlap (Supplementary Figure 2), the corresponding PRS are expected to be correlated. The correlation between the PRS (excluding the *APOE* region) for the four gene-sets is shown in Supplementary Figure 4. The correlation is highest between the oligodendrocyte and synapse gene-sets and is particularly high in AD and PD (corr=0.41 and 0.40, respectively). The next highest correlation is between astrocyte and oligodendrocyte, especially in AD and FTD (corr=0.4 and 0.19, respectively). The correlation between microglia PRS is lowest with other gene-sets across all diseases.

### PRS Association Results

The whole genome PRS (including *APOE*) for each disease is significantly associated with the disease, except for FTD (p=0.323) (Supplementary Table 3). The lack of significant association may be due to limited power as the FTD GWAS has one of the smallest sample sizes and the fewest cases in the UK Biobank.

### Cell-type disease-specific PRS association

Due to the correlation between cell-type specific PRS, we employed Lasso regression modelling to determine the relevance of each cell-type specific PRS with disease, whilst accounting for the other cell-types. In addition, to investigate the degree of the *APOE*-attributed risk in the gene-sets, we jointly modelled the number of *APOE*-e4 and *APOE*-e2 alleles in addition to the cell-type PRS and added PRS excluding all gene-sets and *APOE* (Table 4). Lasso regression is used to select the set of variables that best explain the phenotype. *APOE-*e4 and e2 was included in the joint model to ensure a fair comparison between gene-sets and to highlight which gene-sets are associated beyond the effect of *APOE*. In this setting, the microglia gene-set remains significantly associated with all diseases except for ALS, where only the gene-set containing SNPs not in the four gene-sets or the *APOE* region remains in the final model. For AD, PD with and without dementia and ALS, there is some genetic variation remaining in the PRS excluding all cell-type specific gene-sets. In contrast, for FTD, all variation appears to be explained by *APOE* and microglia.

**Table 4.**
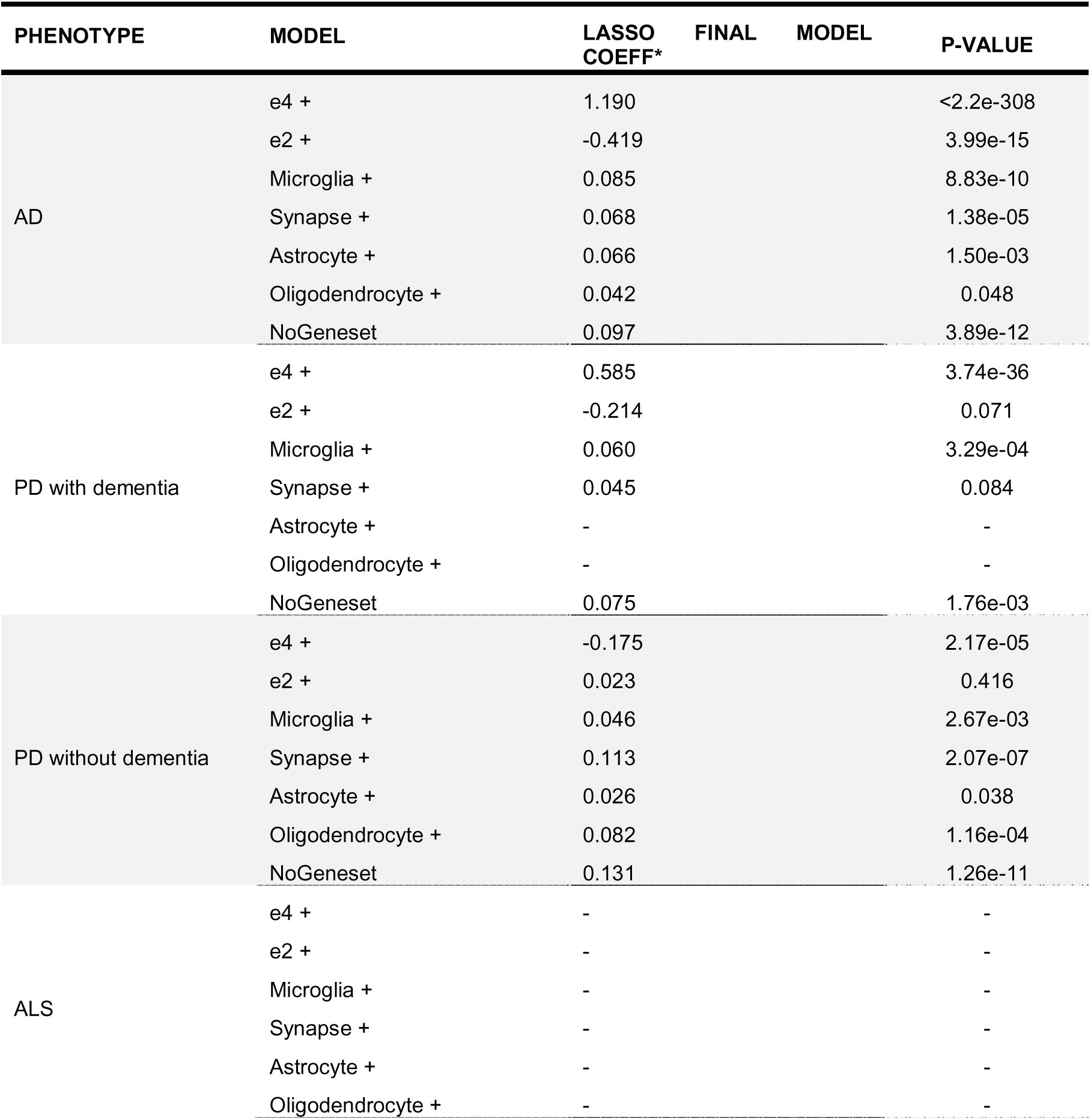
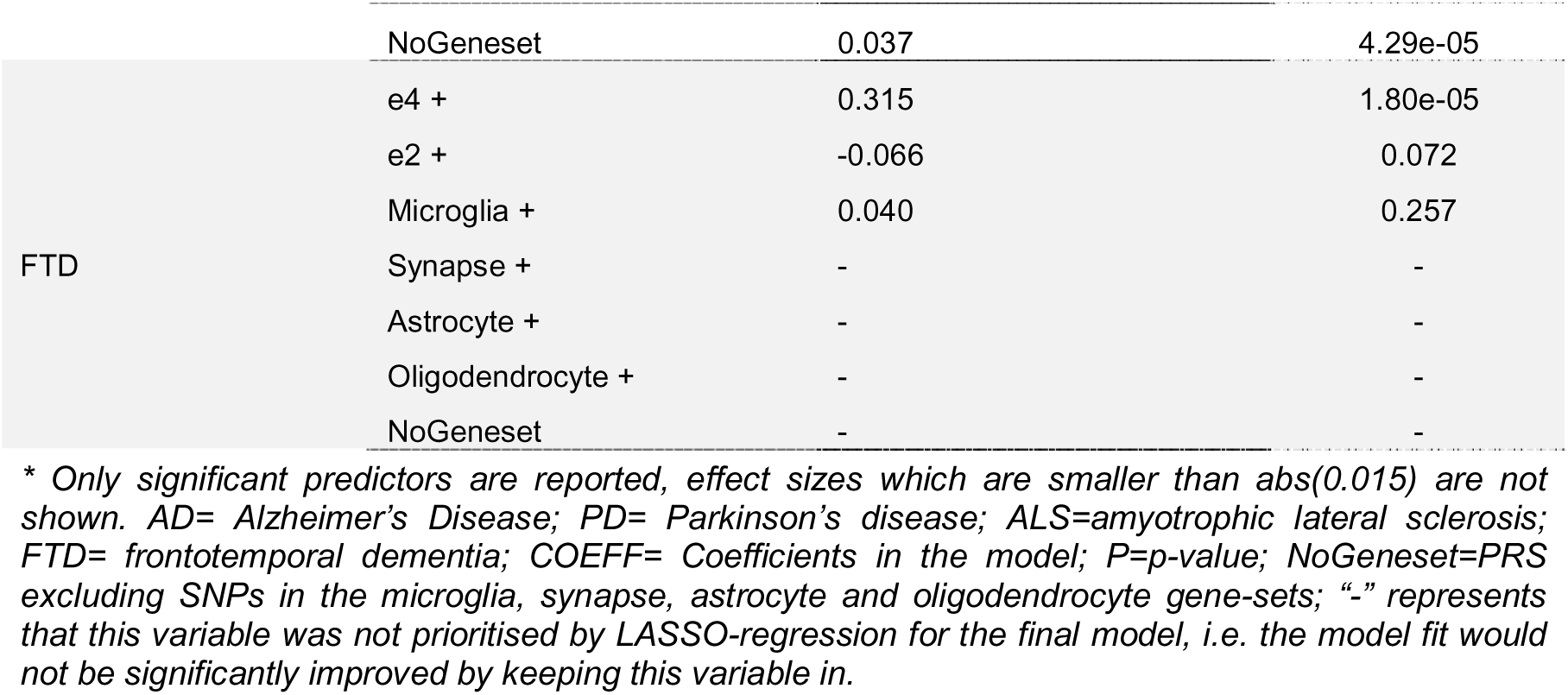
Lasso features selection modelling.

Recognising the interest in disease-specific pathological processes, we present the effects of individual cell type-specific PRSs separately, notwithstanding the substantial overlap among their constituent genes. In these analyses we use Cox regression accounting for age as the cell type-specific genetic liability may capture a specific disease stage which differs by age.

The association analysis between each cell-type disease-specific PRS and the corresponding NDD separately (Supplementary Table 4, full results in Supplementary Table 5), revealed that all gene-sets are significantly associated with AD and PD without dementia with similar effect sizes. As expected, the microglia gene-set has shown a strong association with AD, PD with dementia and PD without dementia (Beta=0.11, SE=0.02, p=1.4e-11, Beta=0.12, SE=0.03, p=3.1e-04, Beta=0.09, SE=0.02, p=6.3e-05, respectively). The strongest association of the synapse gene-set is observed for PD without dementia (Beta=0.16, SE=0.02, p=5.8e-15). The astrocyte gene-set shows a strong association with AD (Beta=0.11, SE=0.02, p=1.6e-11) and a modest association with PD without dementia (Beta=0.06, SE=0.02, p=0.007). The oligodendrocyte gene-set has shown a strong association with AD and PD without dementia (Beta=0.11, SE=0.02, p=1.0e-10, Beta=0.15, SE=0.02, p=1.1e-12, respectively) and a borderline association with ALS (Beta=0.10, SE=0.04, p=0.039).

The similar AUCs observed for the cell-type and whole-genome PRS (Supplementary Table 5) suggest that most of the predictive signal is captured by the gene-set PRS, with limited added value from the whole-genome PRS. Despite the prediction accuracy being similar, it is driven by different components of the PRS and may identify different individuals.

Due to the shared symptoms, misdiagnosis and co-occurrence of neurodegenerative diseases, we also investigated whether a clinically diagnosed NDD has shared genetic loading with other dementias by testing association of the PRS constructed with risk alleles and weights from other NDDs. In this section we have also added vascular dementia (VasDem) as a phenotype in an attempt to use other NDDs and CAD for its genetic prediction. A PRS for VasDem was not assessed in the previous section as there is no large publically available VasDem GWAS that does not include individuals from UKBB. As above we used Cox regression models to determine the impact of PRS including all SNPs, all gene-sets (microglia, synapse, astrocyte and oligodendrocyte as previously defined) and a PRS excluding all gene-sets. Figure 2 shows that all NDDs are significantly associated with all (whole genome and cell-type specific) disease-corresponding PRS, except for FTD (full results can be seen in Supplementary Table 5). Diseases which share dementia as a symptom are predicted by PRS based on multiple diseases. Dementia, AD, vascular dementia and FTD are all associated with AD-, ageing- and DLB-based PRS. PD with dementia is associated with AD-, ageing-, DLB- and PD-based PRS. Only two diseases, PD without dementia and ALS, are only associated with their own disease-corresponding PRS.

**Figure 2.**
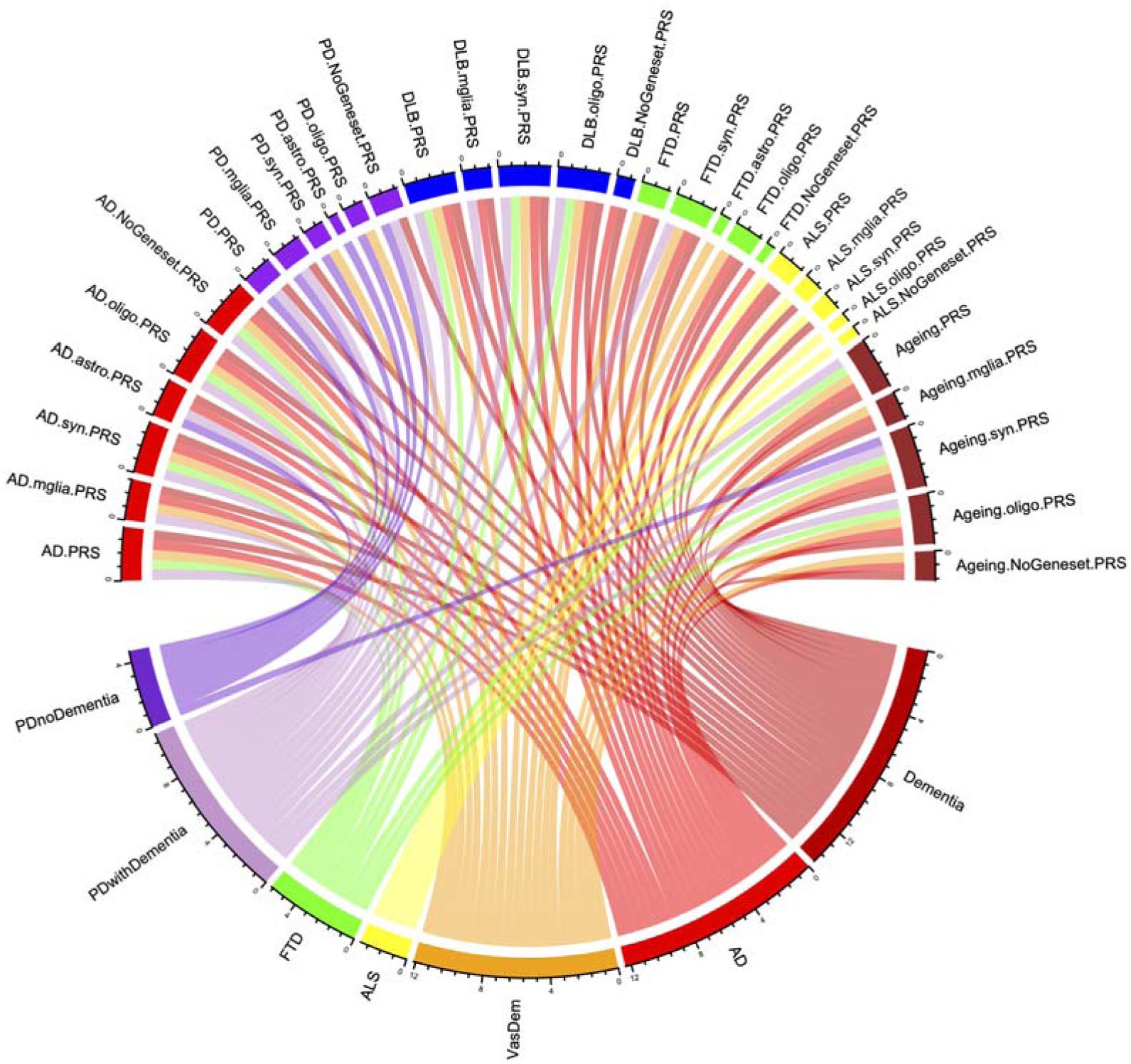
Chord Diagram showing the Prediction Accuracy (AUC at age 70) for all PRS and Neurodegenerative Diseases. Where lines represent a significant association between a PRS and phenotype. These gene-sets include APOE as per gene-set definition.

In addition to AD PRS, other significant contributors to the AD predictions are ageing, DLB and at a lesser strength FTD, especially the microglial, synaptic and oligodendrocyte components (ageing: Beta=-0.49, SE=0.02, p=1.5e-205; DLB: Beta=0.45, SE=0.02, p=2.7e-173; FTD: Beta=0.13, SE=0.02, p=4.9e-14).

PD with dementia, in addition to PD PRS association, also shares genetic liability to AD, ageing and DLB, mostly via microglia, synaptic and oligodendrocyte gene-sets. The prediction accuracy by AD and DLB is similar to that of PD (compare Beta=0.34, SE=0.03, p=2.6e-30 and Beta=0.22, SE=0.03, p=8.9e-14 for AD and DLB, respectively with Beta=0.13, SE=0.03, p=7.8e-05 for PD). In contrast, as mentioned above, PD without dementia is only predicted by PD PRS.

Frontotemporal dementia was better predicted by AD and ageing PRS (AD: Beta=0.30, SE=0.06, p=3.1e-06; Ageing: Beta=-0.15, SE=0.06, p=0.041), as compared to its own FTD PRS (strongest association FTD synapse PRS (Beta=0.14, SE=0.06, p=0.06). ALS was only associated with its own ALS PRS.

Vascular dementia shows the strongest shared genetic loading with ageing and DLB using whole genome-PRS (Beta=-0.33, SE=0.02, p=1.2e-45; Beta=0.25, SE=0.02, p=6.3e-25 for ageing and DLB, respectively), followed by AD (Beta=0.42, SE=0.02, p=2.5e-74). To put it into perspective with other important components of vascular dementia diagnosis and pathogenesis, we tested for the association of VasDem with coronary artery disease (CAD) PRS. We found that whole-genome CAD PRS was strongly associated with vascular dementia (Beta=0.17, SE=0.02, p=1.7e-12), as was CAD microglia PRS (Beta=0.06, SE=0.02, p=0.034), CAD synapse PRS (Beta=0.26, SE=0.02, p=6.8e-27) and CAD oligodendrocyte PRS (Beta=0.10, SE=0.02, p=1.5e-04). CAD PRS was less strongly associated than that of ageing and DLB but with similar prediction accuracy.

### PD with vs without dementia

Since we observe different optimal models in PD with dementia (PDD) and PD without dementia and found a strong association of AD PRS with PDD, we investigated the role of *APOE* in this modelling. From Figure 2 and Supplementary Table 6, AD PRS is more strongly associated with PDD than PD PRS (compare Beta=0.315, SE=0.03, p=1.2e-27 and Beta=0.126, SE=0.03, p=2.3e-05 respectively), and the opposite is true for PD without dementia. **Error! Reference source not found.** Table 4 shows that for PD with and without dementia, the model including both AD and PD PRS shows almost identical coefficients to those when modelled separately, suggesting independent effects of these AD and PD PRS. When *APOE* is included as an additional predictor into the model for PDD, the association with AD PRS is reduced, but remains significant.

Note that when PD without dementia is compared to controls (Table 4), *APOE*-e4 show a protective effect. This protective effect could result from biases such as selection or survival. In terms of selection, our controls were derived from the same source population as the PD patients, based on general elderly population (although healthier than the general UK population ^48^). Survival bias cannot be ruled out, as the age at death of controls is earlier in our sample as compared to PD patients without dementia (see Table 1) and *APOE*-e4 is associated with shortest survival ^49^. Collider bias is quite likely due to selection on “no dementia” status. Furthermore, risk factors for developing dementia in PD include late age of onset for motor manifestation. When looking at PD with dementia in isolation (i.e. comparing to controls without an NDD, there is still a clear association with AD-PRS (equivalent to PD-PRS) in addition to *APOE*.

We also investigated whether and which NDD-corresponding PRS can distinguish between PD with dementia and PD without dementia. Supplementary Table 7 shows the results of these analyses utilising cell-type specific PRS.

Investigating the ability of AD and PD PRS, both separately and together to discriminate between individuals with PD who will/will not develop dementia, we found that the AD PRS is strongly associated with the difference between PD with and without dementia (Beta=0.362, SE=0.036, p=3.2e-23, Table 5) with most prediction accuracy explained by the joint effects of the AD and PD PRS (compare R2=0.038 for that the AD PRS, Table 5, R2=0.04 for the joint effects). The inclusion of *APOE*-e4 in the model explains most of the AD PRS association. *APOE*-e4 alone separates PD individuals with vs without dementia with R2=0.061. AD and PD PRS remain significant predictors over and above *APOE*-e4, increasing the R2=0.064.

**Table 5.** NDD vs NDD analyses.

| PHENOTYPE | MODEL | COEFF | BETA | SE | P | R2 | AUC |
| --- | --- | --- | --- | --- | --- | --- | --- |
| PD with vs. PD without dementia | PD PRS |  | -0.082 | 0.036 | 0.023 | 0.002 | 0.52 |
|  | AD PRS |  | 0.362 | 0.036 | 3.2e-23 | 0.038 | 0.60 |
|  | PD PRS + AD PRS | PD PRS | -0.077 | 0.036 | 0.034 | 0.040 | 0.60 |
|  |  | AD PRS | 0.360 | 0.036 | 4.4e-23 |  |  |
|  | e4 | e4 | 0.837 | 0.066 | 1.6e-36 | 0.061 | 0.61 |
|  | e4 + PD PRS + AD PRS | e4 | 0.702 | 0.087 | 7.7e-16 | 0.064 | 0.62 |
|  |  | PD PRS | -0.078 | 0.037 | 0.034 |  |  |
|  |  | AD PRS | 0.112 | 0.047 | 0.018 |  |  |

*AD= Alzheimer's Disease; PD= Parkinson's disease; COEFF= Coefficients in the model; BETA= log odds ratio from the model; SE=standard error; P=p-value; R2=the amount of variability in the phenotype explained by the model.*

We also investigate the other NDDs with known evidence of shared genetics, such as ALS and FTD, or AD and FTD, results are as expected; *APOE* only separates both ALS and FTD and AD and FTD, see Supplementary Tables 7 and 8.

### Gene-set polygenic risk scores and Biomarkers

Finally, we investigated the association between neurofilament light (NfL) and glial fibrillary acidic protein (GFAP) with gene-set PRS for all NDDs, see Supplementary Figure 5, here we used gene-sets as defined by the publication (i.e. including *APOE* as per gene-set definition). The microglia gene-set is additionally filtered to include established regulatory regions of the genes, see Supplementary Methods. The association between gene-set PRS and GFAP were in general much stronger than those with NfL. In AD, ageing and DLB, the synapse gene-set was associated with NfL (Beta=0.01, SE=0.003, p=1.9e-04; Beta=-0.01, SE=0.003, p=2.5e-07; and Beta=0.01, SE=0.003, p=7.0e-04, respectively), and in AD the oligodendrocyte gene-set was associated with NfL (Beta=0.009, SE=0.003, p=7.0e-04). For GFAP there was an association between microglia, synapse and oligodendrocyte gene-set PRS for AD (Beta=0.01, SE=0.003, p=5.7e-06; Beta=0.03, SE=0.003, p=6.1e-25 and Beta=0.03, SE=0.003, p=2.7e-24), whereas the synapse gene-set was associated with ageing, DLB and FTD (Beta=-0.03, SE=0.003, p=9.2e-35; Beta=0.03, SE=0.003, p=5.1e-28; and Beta=0.01, SE=0.003, p=8.2e-05, respectively) and the oligodendrocyte gene-set was associated with GFAP for ageing and DLB (Beta=-0.01, SE=0.003, p=1.1e-06 and Beta=0.03, SE=0.003, p=4.2e-22, respectively). Note that GFAP was not significantly associated with any NDD astrocyte PRS.

## Discussion

In this study we draw together data from all large NDD genome wide association studies for AD, PD, ALS, FTD and investigate PRS predictive ability with the aim to understand and quantify the mixed pathology in UKB individuals, who have been diagnosed with one or multiple NDDs. For this, we derived both whole-genome and cell-type specific PRS for each NDD and tested their ability to discriminate between NDDs and healthy controls using disease-corresponding PRS and cross-disease PRS. We have also attempted to discriminate between PD with and without dementia.

We reach the conclusion that the prediction accuracy of NDDs using cell-type specific PRS is the same or higher than that of whole-genome PRS, although this prediction accuracy remains insufficient for clinical utility. However, since the gene-set PRS contains a much smaller number of SNPs than the whole-genome PRS and reflects biological mechanisms underlying disease development and progression, the interpretation of the results is more transparent, and the uncertainty associated with an individual’s PRS is likely to be reduced compared with the whole-genome approach ^50^.

The results of this study show that variants in genes expressed in all gene-sets (microglia, synapses, astrocytes and oligodendrocytes) are associated with AD over and above *APOE* with the microglial gene-set remaining the most strongly associated when analysed accounting for other cell-types. This result fits with a wealth of cellular studies in model organisms and with postmortem human tissue. Microglia are important in AD as they play a role in maintaining neural homeostasis and responding to injury or disease ^15^. *APOE* is upregulated more in microglia than other cell types in AD. Synaptic dysfunction plays a pivotal role in the progression of AD, with growing evidence suggesting it is a key driver of cognitive decline ^19^. Synapse loss is one of the strongest correlates of cognitive impairment in AD and often precedes overt neuronal death ^51^. The pathological hallmarks of AD, amyloid-beta (Aβ) plaques and tau tangles, disrupt synaptic function through multiple mechanisms, including impairing synaptic plasticity and altering neurotransmission ^51,52^. Astrocytes are known to be important in AD, as they are essential for maintaining neuronal health ^20^. Oligodendrocytes also show functional changes in AD ^24^ and contribute to the development of pathology ^25,26^. In addition, studies have shown that different cell types influence distinct stages of AD progression ^53^. Synaptic dysfunction is strongly associated with the early stages of AD, particularly during mild cognitive impairment, as evidenced by alterations in CSF synaptic proteins during these stages ^54^. Oligodendrocytes, which are responsible for myelin production, are also affected very early in AD progression ^55^. The relatively young age of the participants in our study (mean age ∼74 years) suggests that the AD cases were at an early stage of disease progression, with most individuals being within approximately five years of diagnosis (Supplementary Figure 6). This may further explain why the predictive performance of these gene sets was comparable to that of the well-established microglial gene set in our study.

PD without dementia is explained by microglia, synapse, astrocyte and oligodendrocyte gene-sets, ALS is explained by the gene-set excluding all cell-type genes and *APOE*, PD with dementia is explained by microglia, synapse and astrocyte, and FTD is explained by microglia only. It was observed that PD with dementia was mostly attributed to the microglia cell-type despite it previously being suggested that PD genes are related to synaptic homeostasis ^56^ and early synaptic dysfunction is a hallmark of PD, even before cognitive symptoms appear. However, typically PD GWAS studies combine PD with/without dementia. In PD patients without dementia, synaptic markers such as synaptophysin and SV2A are reduced in cortical regions like the anterior cingulate and insula ^57^. In contrast, dementia in PD is not solely driven by synaptic loss, but by other co-pathologies such as inflammation ^58^, alpha-synuclein accumulation and amyloid-beta and tau pathology (also seen in AD) ^59^, which may overshadow the role of synapses in PD with dementia. The difference in findings for PD with and without dementia is likely due to PD with dementia being a distinct phase of PD ^60^.

For PD with dementia the microglial gene set exhibited the strongest association with disease risk compared with the other cell-type-specific gene sets. The association between microglia and PD is reported in the literature with an association between the microglia proportion and type 1 interferon response in early PD ^61^. Across the other disorders the prediction accuracy is relatively consistent across all gene-sets. Once the SNPs within the cell-type specific gene-sets and *APOE* are removed, there is no remaining genetic association for FTD (although this may be caused by small sample size or misdiagnosis), whereas other NDDs explain additional genetic variation beyond the cell-type specific gene-sets.

In general, diseases which exhibit dementia symptoms suggest shared pathology, mechanisms and/or disease aetiology between them ^62^. Our study revealed frequent co-occurring diagnoses, particularly involving AD, vascular dementia, PDD, and FTD. We also show a substantial overlap in PRS prediction, especially where dementia is a symptom, which is likely to be due to shared disease aetiology and/or shared genetic variants across diseases. Across neurodegenerative diseases, AD predictions are influenced not only by AD PRS but also strongly by ageing, DLB, and to a lesser extent FTD, with shared effects largely driven by microglial, synaptic, and oligodendrocyte pathways. PD with dementia shows additional shared genetic liability with AD, ageing, and DLB, and is predicted with similar accuracy by AD and DLB risk scores as by PD risk scores, whereas PD without dementia is mainly driven by its own genetic risk. Vascular dementia shows the strongest overlap with ageing and DLB, followed by AD and coronary artery disease genetics, indicating broad shared biological contributions across neurodegeneration and vascular pathology, while ALS remains primarily associated with its own genetic risk score.

Looking at biomarkers for neurodegeneration, synapse PRS was associated with GFAP and NfL for AD, ageing and DLB and oligodendrocyte PRS was associated with GFAP for AD, ageing and DLB. Surprisingly, GFAP was not significantly associated with any of NDD astrocyte PRS, implying that GFAP as a biomarker may reflect dysfunction of astrocytes after other cells, i.e. astrocytes may be responding to malfunctions of surrounding cells. While GFAP is commonly used as a marker of astrocytes, particularly RNA-sequencing experiments have shown that *GFAP* is also expressed by non-traditional astrocytes or cell types that appear to have complex identities, often at lower expression levels ^63^.

A major strength of this study is the use of UK Biobank data, which offers rich genetic and phenotypic information. Applying the same analytical framework across multiple cell types within a single dataset allowed us to investigate cell-type-specific PRSs across several neurodegenerative disorders in a directly comparable manner. Leveraging the large sample size, we were able to model the association of each cell-type-specific PRS while adjusting for the effects of the other cell types and *APOE*. Due to time-stamped data in UK Biobank, we were able to use Cox models when modelling each cell-type PRS separately, which account for age, which is a substantial confounder in neurodegenerative disease, and to also account for age at death. This is particularly important in cross-disorder analyses when age at onset differs between NDDs. The extensive phenotype information has allowed for differentiation between PD with and without dementia, which have clearly shown different genetic components, and in most studies would have been grouped together.

We would be remiss if we did not mention limitations of the study. First, there is always the issue of misdiagnosis with neurodegenerative diseases, since there are many overlapping symptoms and it is difficult to differentiate between secondary and revised diagnoses in UK Biobank. The diagnosis accuracy in UK Biobank has been assessed previously and the diagnosis of all cause dementia and AD were reasonable, but predictive value was lower for other dementia subtypes ^64^.

Second, the imprecision of disease-specific SNP effect size estimates will inevitably lead to less precise estimation of genetic liability by PRS, particularly as the number of SNPs included in the PRS increases ^50^. The quality of SNP effect size estimates depends on increasing GWAS sample size while preserving the precision and reliability of disease diagnosis, as well as the use of age-matched controls. In this study, we were only able to compute PRS for NDDs with available summary statistics that did not include UK Biobank participants. In addition, the summary statistics for FTD and DLB were derived from considerably smaller sample sizes than those available for other NDDs, which may reduce the predictive power of their corresponding PRS. Furthermore, the number of individuals with specific NDDs in the UK Biobank varies substantially, meaning that the statistical association results are not directly comparable in terms of significance.

Third, we used cell-type specific gene sets derived from published data; however, more predictive unpublished gene sets may exist that could provide improved comparability. Nevertheless, we aimed to leverage the most robust and well-validated gene-set information currently available in the public domain. In addition, the gene-sets were not defined uniformly across studies. This may have reduced the precision and statistical power of gene-sets that have been less extensively studied in the context of NDDs and are therefore less well characterised. The number of SNPs included in each gene-set also varied considerably, which may both increase the amount of captured genetic liability and introduce additional noise. Despite these differences in SNP counts across gene sets, we observed comparable and biologically plausible contributions of all cell types to AD prediction, which provides reassurance regarding the validity of the findings for other NDDs. The cell-type specific gene-sets showed substantial SNP overlap, since the gene-sets are derived independently by separate research groups without considering other cell-types. In addition, some genes are expected to have multiple functions and so are present in multiple gene-sets, however the top terms highlighted by gprofiler for the gene-sets are different, suggesting that they do capture different processes.

Finally, the participation bias and the age distribution of UK Biobank participants limit our analyses to estimating cell-type specific association significance and accuracy only within this specific context and may lead to over- or underestimation of association strengths in datasets with different age distributions or study designs.

In summary, this study provides important conceptual groundwork in the field of dementia by supporting the idea that future therapeutic strategies will need to target specific brain cell-types. It also highlights the limitations of current clinical diagnostic practices and the challenges faced by biological research in identifying truly cell-type specific genes. To elucidate disease stage-specific genetic signatures, large, longitudinal, population-based studies are required. To conclude, understanding the relationship between genetic risk, cell types, and therapeutic targets is likely to be central to treatment success.

## Author Contributions

ES - analysed the data, drafted and reviewed the manuscript; AMK, GL, PT, SC, MR, BK, JOB, TSJ - contributed to interpretation of data and critically reviewed the manuscript; UY, DAS – curated the oligodendrocyte gene-list, contributed to interpretation of data and critically reviewed the manuscript; KK - designed and created Figures; VEP - participated in conception and design of the work, analysed and interpreted the data, drafted and reviewed the manuscript. All authors approved the final version.

## Data Availability

Access to UK Biobank data can be requested from https://www.ukbiobank.ac.uk/use-our-data/apply-for-access/. All polygenic risk scores, whole genome and cell type specific, and SNP lists for each cell type will be returned to UK Biobank and made available to other researchers using UK Biobank.

## Acknowledgements

This research has been conducted using the UK Biobank Resource under application number 109607. We would like to acknowledge Dr Francesco Fiorini for his initial discussion about co-occurring diagnoses in dementia.

