## Supplementary material for "Mapping polygenic risk scores to brain cells: insights for neurodegenerative diseases": SupplementaryMaterial.docx

Supplementary Table 1- **Number of Co-occurring Dementia Diagnoses in UK Biobank.** Where AD=Alzheimer’s Disease, VasDem=Vascular Dementia, ALS=Amyotrophic Lateral Sclerosis, FTD=Frontotemporal Dementia, PD_dem=Parkinson’s Disease with dementia, PD_nodem= Parkinson’s Disease without dementia.

| **Diagnosis 1** | **Diagnosis 2** | **Number of co-occurring diagnoses (% diagnosis 1, % diagnosis 2)** |
| --- | --- | --- |
| AD (N=3,729) | VasDem (N=1,805) | 426 (11.4, 23.6) |
| AD (N=3,729) | ALS (N=631) | 10 (0.3, 1.6) |
| AD (N=3,729) | FTD (N=261) | 46 (1.2, 17.6) |
| AD (N=3,729) | PD_dem (N=1,132) | 251 (6.7, 22.2) |
| AD (N=3,729) | PD_nodem (N=2,549) | 0 |
| VasDem (N=1,805) | ALS (N=631) | 4 (0.2, 0.6) |
| VasDem (N=1,805) | FTD (N=261) | 27 (1.5, 10.3) |
| VasDem (N=1,805) | PD_dem (N=1,132) | 152 (8.4, 13.4) |
| VasDem (N=1,805) | PD_nodem (N=2,549) | 0 |
| ALS (N=631) | FTD (N=261) | 17 (2.7, 6.5) |
| ALS (N=631) | PD_dem (N=1,132) | 5 (0.8, 0.4) |
| ALS (N=631) | PD_nodem (N=2,549) | 21 (3.3, 0.8) |
| FTD (N=261) | PD_dem (N=1,132) | 17 (6.5, 1.5) |
| FTD (N=261) | PD_nodem (N=2,549) | 0 |
| PD_dem (N=1,132) | PD_nodem (N=2,549) | 0 |

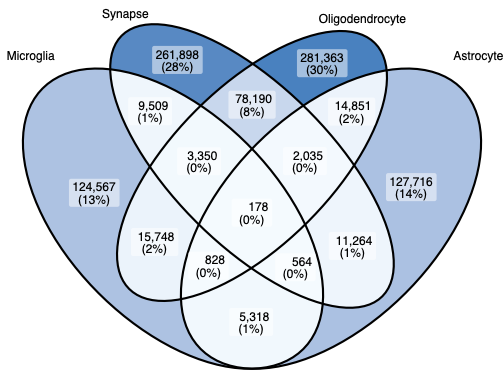

Supplementary Figure 1- **Venn Diagram showing the overlap between SNPs in cell specific gene-sets; microglia, synapse, oligodendrocyte and astrocyte, with APOE included as per gene-set definition.** Each oval represents the SNPs in each cell-type specific gene-set and the overlap shows the number of SNPs overlapping between these gene-sets, and the percentage is the percentage of the total number of SNPs across all gene-sets.

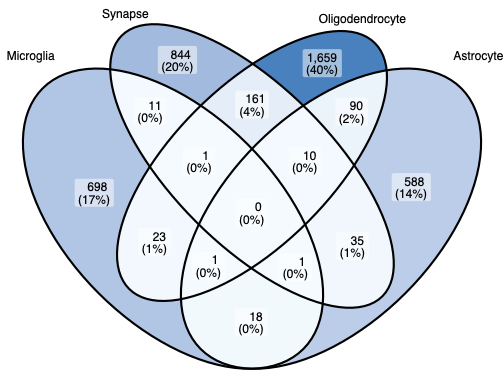

Supplementary Figure 2- Venn Diagram showing the overlap between genes in cell specific gene-sets, with APOE included as per gene-set definition

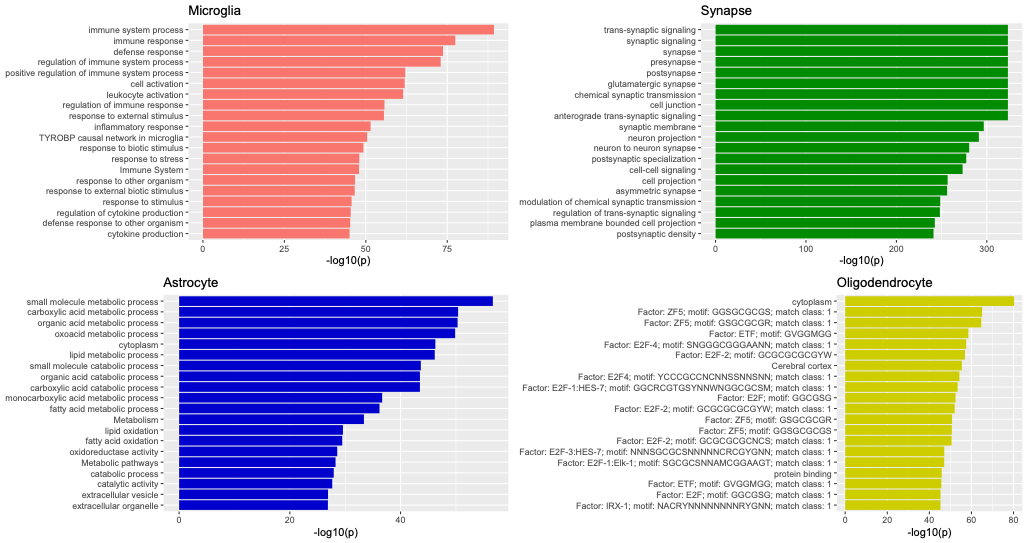

Supplementary Figure 3- Gene enrichment analysis for the cell type specific gene-sets

Supplementary Table 2 - AUC Comparison for Endo et al. and Mathys et al. defined astrocyte gene-sets

| **Disease** | **AUC timepoint** | **Endo – Mean (SD)** | **Mathys – Mean (SD)** |
| --- | --- | --- | --- |
| All NDDs | 60 | 0.50 (0.03) | 0.51 (0.03) |
|  | 70 | 0.50 (0.02) | 0.51 (0.02) |
|  | 80 | 0.51 (0.01) | 0.51 (0.01) |
| AD | 60 | 0.49 (0.05) | 0.51 (0.03) |
|  | 70 | 0.51 (0.02) | 0.53 (0.03) |
|  | 80 | 0.51 (0.01) | 0.53 (0.01) |
| PD | 60 | 0.51 (0.02) | 0.50 (0.04) |
|  | 70 | 0.49 (0.01) | 0.52 (0.02) |
|  | 80 | 0.51 (0.01) | 0.52 (0.01) |
| DLB | 60 | 0.50 (0.02) | 0.51 (0.02) |
|  | 70 | 0.51 (0.02) | 0.51 (0.01) |
|  | 80 | 0.50 (0.005) | 0.51 (0.008) |
| FTD | 60 | 0.51 (0.05) | 0.48 (0.03) |
|  | 70 | 0.50 (0.01) | 0.51 (0.01) |
|  | 80 | 0.51 (0.01) | 0.51 (0.005) |
| ALS | 60 | 0.50 (0.02) | 0.53 (0.03) |
|  | 70 | 0.50 (0.02) | 0.50 (0.01) |
|  | 80 | 0.50 (0.005) | 0.51 (0) |
| Ageing | 60 | 0.50 (0.02) | 0.51 (0.02) |
|  | 70 | 0.50 (0.007) | 0.50 (0.02) |
|  | 80 | 0.51 (0.005) | 0.51 (0.008) |

*AUC= area under the curve; SD=standard deviation; NDD=neurodegenerative disease; AD= Alzheimer’s Disease; PD= Parkinson’s disease; VasDem= vascular dementia; ALS=amyotrophic lateral sclerosis; FTD= frontotemporal dementia.*

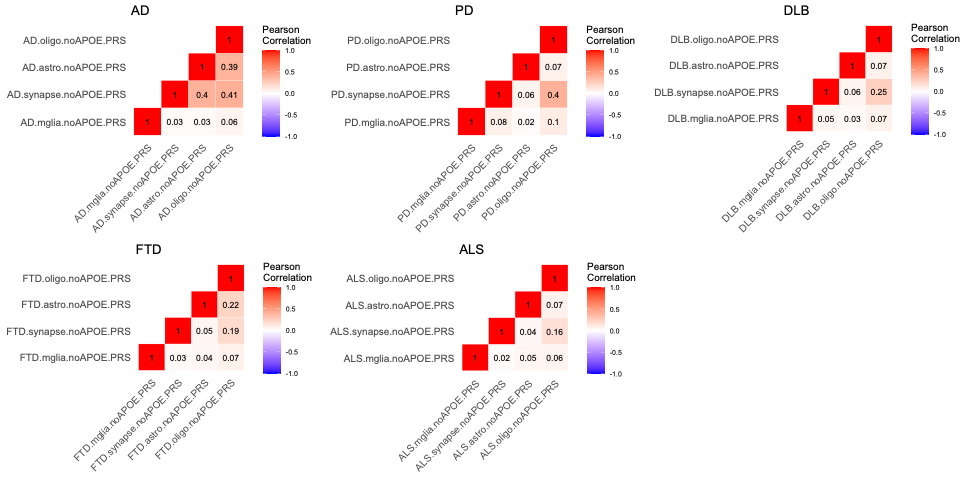

Supplementary Figure 4- Correlation Heatmaps between gene-set PRS in each neurodegenerative disorder

Supplementary Table 3- Association and Prediction Accuracy for NDDs with disease corresponding whole genome PRS.

| PHENOTYPE | MODEL | HR | SE | Padj | R2 | AUC at age 70 |
| --- | --- | --- | --- | --- | --- | --- |
| AD | AD-PRS | 1.93 | 0.02 | <2.2e-308 | 0.369 | 0.67 |
| ALS | ALS-PRS | 1.21 | 0.04 | 9.5e-06 | 0.081 | 0.59 |
| FTD | FTD-PRS | 1.08 | 0.06 | 0.323 | 0.062 | 0.54 |
| PD with Dementia | PD-PRS | 1.14 | 0.03 | 7.4e-05 | 0.187 | 0.65 |
| PD no Dementia | PD-PRS | 1.23 | 0.02 | 6.8e-25 | 0.126 | 0.60 |

*AD= Alzheimer’s Disease; PD= Parkinson’s disease; ALS=amyotrophic lateral sclerosis; FTD= frontotemporal dementia; PRS=polygenic risk score; COEFF= Coefficients in the model; HR= hazards ratio; SE=standard error; Padj=FDR adjusted p-value; R2=the proportion of variability in the phenotype explained by the model; AUC=area under the curve.*

Supplementary Table 4- Association and Prediction Accuracy of NDD with cell-type-specific PRS, constructed with disease-corresponding GWAS SNP effects

| PHENOTYPE | MODEL | HR | SE | Padj | R2 | AUC at age 70 | AUC with APOE at age 70 |
| --- | --- | --- | --- | --- | --- | --- | --- |
| AD | AD.mglia.PRS | 1.12 | 0.02 | 1.4e-11 | 0.01 | 0.53 | 0.69 |
|  | AD.synapse.PRS | 1.14 | 0.02 | 5.5e-15 | 0.02 | 0.56 | 0.70 |
|  | AD.astrocyte.PRS | 1.12 | 0.02 | 1.6e-11 | 0.01 | 0.54 | 0.70 |
|  | AD.oligodendrocyte.PRS | 1.12 | 0.02 | 1.0e-10 | 0.01 | 0.53 | 0.69 |
| ALS | ALS.mglia.PRS | 1.09 | 0.04 | 0.084 | 0.06 | 0.55 | 0.55 |
|  | ALS.synapse.PRS | 1.13 | 0.04 | 0.007 | 0.06 | 0.57 | 0.57 |
|  | ALS.astrocyte.PRS | 0.97 | 0.04 | 0.570 | 0.05 | 0.56 | 0.56 |
|  | ALS.oligodendrocyte.PRS | 1.11 | 0.04 | 0.039 | 0.06 | 0.57 | 0.57 |
| FTD | FTD.mglia.PRS | 1.07 | 0.06 | 0.414 | 0.06 | 0.57 | 0.58 |
|  | FTD.synapse.PRS | 1.04 | 0.06 | 0.669 | 0.06 | 0.57 | 0.58 |
|  | FTD.astrocyte.PRS | 1.04 | 0.06 | 0.682 | 0.06 | 0.56 | 0.58 |
|  | FTD.oligodendrocyte.PRS | 1.05 | 0.06 | 0.583 | 0.06 | 0.55 | 0.57 |
| PD with dementia | PD.mglia.PRS | 1.13 | 0.03 | 3.1e-04 | 0.19 | 0.63 | 0.66 |
|  | PD.synapse.PRS | 1.07 | 0.03 | 0.07 | 0.18 | 0.64 | 0.67 |
|  | PD.astrocyte.PRS | 1.00 | 0.03 | 0.995 | 0.17 | 0.62 | 0.65 |
|  | PD.oligodendrocyte.PRS | 1.06 | 0.03 | 0.121 | 0.18 | 0.63 | 0.66 |
| PD no dementia | PD.mglia.PRS | 1.09 | 0.02 | 6.3e-05 | 0.09 | 0.58 | 0.59 |
|  | PD.synapse.PRS | 1.17 | 0.02 | 5.8e-15 | 0.11 | 0.59 | 0.59 |
|  | PD.astrocyte.PRS | 1.06 | 0.02 | 0.007 | 0.09 | 0.58 | 0.58 |
|  | PD.oligodendrocyte.PRS | 1.16 | 0.02 | 1.1e-12 | 0.11 | 0.59 | 0.59 |

*Note that all gene-sets exclude the APOE region (chr19:44.4-46.5Mb, hg19, and correspond to .noAPOE. results in Supplementary Table 2. AD= Alzheimer’s Disease; PD= Parkinson’s disease; ALS=amyotrophic lateral sclerosis; FTD= frontotemporal dementia; PRS=polygenic risk score; COEFF= Coefficients in the model; HR= hazard ratio; SE=standard error; Padj=FDR adjusted p-value; R2=the amount of variability in the phenotype explained by the model; AUC=area under the curve; mglia= microglia.*

Supplementary Table 5- Association and Prediction Accuracy for Cox Proportional Hazards Models Between All PRSes and Neurodegenerative Disorders

(excel sheet)

*NPheno= Number of individuals with that phenotype; PRS= polygenic risk score; HR=hazard ratio; SE=standard error; P= unadjusted p-value; Padj= FDR corrected p-value; AUC60= area under the curve at age 60; AUC70= area under the curve at age 70; AUC80= area under the curve at age 80; R2 is the amount of variability in the phenotype explained by the PRS; AD= Alzheimer’s Disease; PD= Parkinson’s disease; VasDem= vascular dementia; ALS=amyotrophic lateral sclerosis; FTD= frontotemporal dementia.*

Supplementary Table 6- PRS Models for Parkinson's Disease

| PHENOTYPE | MODEL | COEFF | BETA | SE | P | R2 | AUC |
| --- | --- | --- | --- | --- | --- | --- | --- |
| PD with dementia (vs. controls) | PD PRS |  | 0.126 | 0.030 | 2.3e-05 | 0.0012 | 0.54 |
|  | AD PRS |  | 0.315 | 0.029 | 1.2e-27 | 0.008 | 0.59 |
|  | PD PRS + AD PRS | PD PRS  AD PRS | 0.126  0.315 | 0.030  0.029 | 2.1e-05  1.1e-27 | 0.009 | 0.60 |
|  | e4 | e4 | 0.627 | 0.048 | 1.5e-38 | 0.010 | 0.59 |
|  | e4 + PD PRS + AD PRS | e4  PD PRS  AD PRS | 0.465  0.127  0.134 | 0.068  0.030  0.039 | 5.5e-12  1.9e-05  5.9e-04 | 0.012 | 0.61 |
| PD without dementia (vs. controls) | PD PRS |  | 0.208 | 0.020 | 1.0e-25 | 0.004 | 0.56 |
|  | AD PRS |  | -0.039 | 0.020 | 0.052 | 0.0001 | 0.51 |
|  | PD PRS + AD PRS | PD PRS  AD PRS | 0.208  -0.038 | 0.020  0.020 | 1.0e-25  0.054 | 0.004 | 0.56 |
|  | e4 | e4 | -0.181* | 0.041 | 1.2e-05 | 0.0007 | 0.52 |
|  | e4 + PD PRS + AD PRS | e4  PD PRS  AD PRS | -0.211  0.208  0.026 | 0.052  0.020  0.025 | 4.3e-05  1.2e-25  0.305 | 0.004 | 0.56 |

** The negative association is likely driven by the collider bias (see discussion).*

Supplementary Table 7- NDD vs NDD analyses

| PHENOTYPE | MODEL | COEFF | BETA | SE | P | R2 | AUC |
| --- | --- | --- | --- | --- | --- | --- | --- |
| PD with vs. PD without dementia | AD.mglia + AD.syn + AD.ast +  AD.oligo | Mglia  Synapse  Astrocyte  Oligo | 0.048  0.012  -0.005  0.054 | 0.036  0.040  0.040  0.040 | 0.179  0.772  0.897  0.184 | 0.002 | 0.52 |
|  | PD.mglia + PD.syn + PD.ast +  PD.oligo | Mglia  Synapse  Astrocyte  Oligo | 0.046  -0.073  -0.054  -0.068 | 0.036  0.039  0.036  0.039 | 0.207  0.062  0.132  0.083 | 0.005 | 0.54 |
| ALS vs FTD | ALS.mglia + ALS.syn + ALS.ast +  ALS.oligo | Mglia  Synapse  Astrocyte  Oligo | 0.161  -0.037  -0.010  0.124 | 0.074  0.074  0.073  0.073 | 0.030  0.620  0.896  0.088 | 0.013 | 0.56 |
|  | FTD.mglia + FTD.syn + FTD.ast +  FTD.oligo | Mglia  Synapse  Astrocyte  Oligo | 0.053  -0.074  -0.022  -0.092 | 0.076  0.079  0.077  0.076 | 0.488  0.344  0.775  0.226 | 0.006 | 0.54 |
| AD vs Vascular dementia | AD.mglia + AD.syn + AD.ast +  AD.oligo | Mglia  Synapse  Astrocyte  Oligo | 0.074  0.010  0.089  -0.054 | 0.029  0.033  0.033  0.034 | 0.011  0.763  0.007  0.108 | 0.004 | 0.53 |
| AD vs FTD | AD.mglia + AD.syn + AD.ast +  AD.oligo | Mglia  Synapse  Astrocyte  Oligo | -0.011  0.013  0.099  -0.065 | 0.064  0.072  0.072  0.074 | 0.861  0.862  0.171  0.374 | 0.002 | 0.53 |
|  | FTD.mglia + FTD.syn + FTD.ast +  FTD.oligo | Mglia  Synapse  Astrocyte  Oligo | -0.063  -0.030  -0.020  -0.025 | 0.065  0.066  0.066  0.067 | 0.337  0.645  0.760  0.709 | 0.001 | 0.52 |

*Note that these gene-sets do not include the APOE region (chr19: 44.4-46.5Mb, hg19). AD= Alzheimer’s Disease; PD= Parkinson’s disease; COEFF= Coefficients in the model; BETA= log odds ratio from the model; SE=standard error; P=p-value; R2=the amount of variability in the phenotype explained by the model.*

Supplementary Table 8- NDD vs NDD analyses

| PHENOTYPE | MODEL | COEFF | BETA | SE | P | R2 | AUC |
| --- | --- | --- | --- | --- | --- | --- | --- |
| ALS vs FTD | ALS PRS |  | 0.059 | 0.071 | 0.401 | 0.001 | 0.49 |
|  | FTD PRS |  | -0.061 | 0.073 | 0.409 | 0.001 | 0.49 |
|  | ALS PRS + FTD PRS | ALS PRS  FTD PRS | 0.063  -0.064 | 0.071  0.074 | 0.376  0.384 | 0.002 | 0.52 |
|  | e4 | e4 | -0.555 | 0.132 | 2.6e-05 | 0.028 | 0.56 |
|  | e4 + ALS PRS + FTD PRS | e4  ALS PRS  FTD PRS | -0.547  0.057  -0.030 | 0.133  0.072  0.075 | 3.9e-05  0.426  0.687 | 0.029 | 0.57 |
| AD vs Vascular dementia | AD PRS |  | 0.236 | 0.027 | 2.9e-18 | 0.021 | 0.58 |
|  | e4 | e4 | 0.437 | 0.045 | 4.5e-22 | 0.026 | 0.58 |
|  | e4 + AD PRS | e4  AD PRS | 0.323  0.094 | 0.065  0.039 | 6.7e-07  0.016 | 0.027 | 0.58 |
| AD vs FTD | AD PRS |  | 0.311 | 0.058 | 9.9e-08 | 0.019 | 0.61 |
|  | FTD PRS |  | 0.048 | 0.064 | 0.456 | 0.0004 | 0.52 |
|  | AD PRS + FTD PRS | AD PRS  FTD PRS | 0.310  0.012 | 0.059  0.064 | 1.3e-07  0.848 | 0.019 | 0.61 |
|  | e4 | e4 | 0.743 | 0.108 | 6.9e-12 | 0.034 | 0.62 |
|  | e4 + AD PRS + FTD PRS | e4  AD PRS  FTD PRS | 0.717  0.021  -0.002 | 0.149  0.084  0.065 | 1.5e-06  0.797  0.980 | 0.034 | 0.63 |

*AD= Alzheimer’s Disease; PD= Parkinson’s disease; COEFF= Coefficients in the model; BETA= log odds ratio from the model; SE=standard error; P=p-value; R2=the amount of variability in the phenotype explained by the model.*

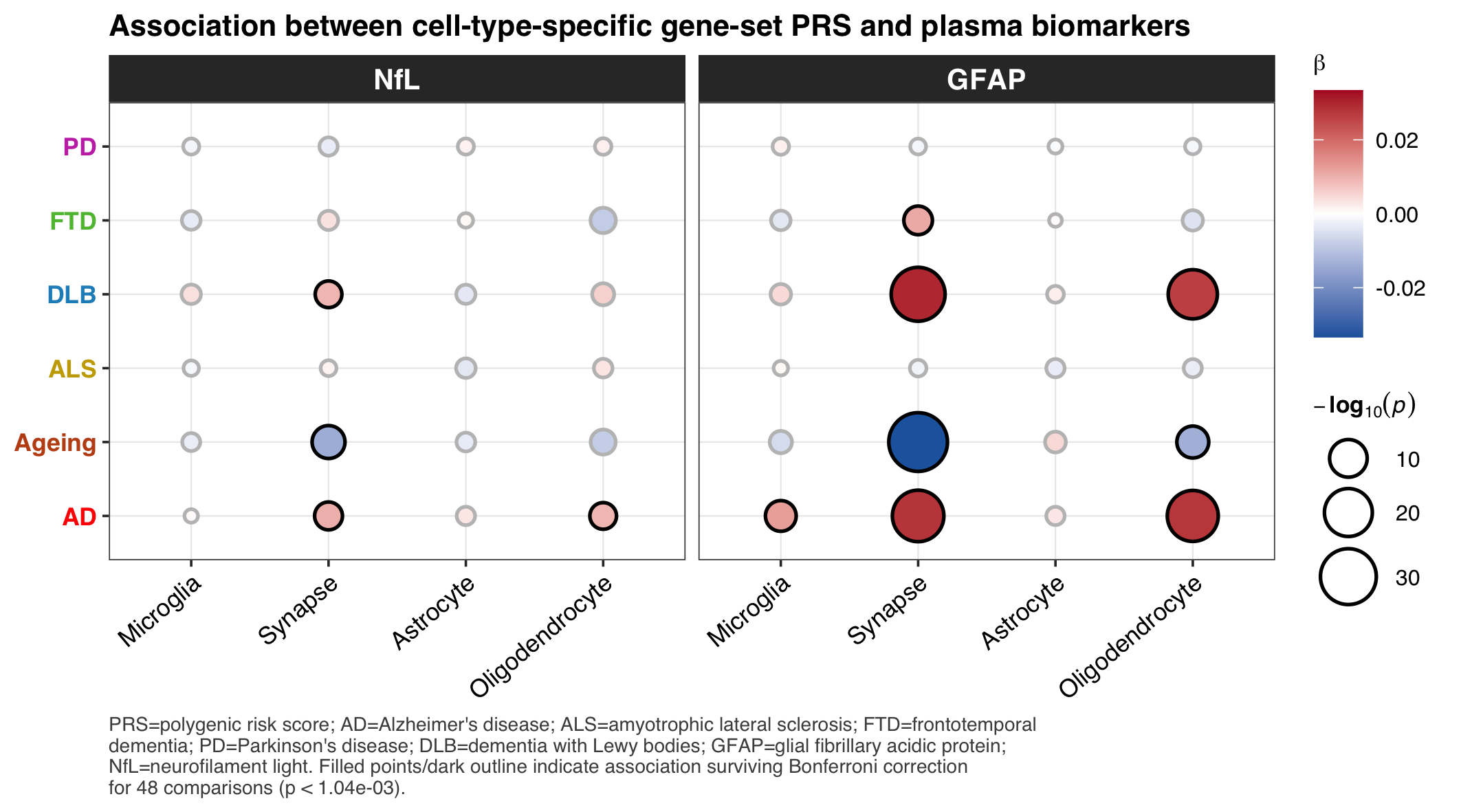

Supplementary Figure 5- **Association Between Gene-set PRS and Biomarkers (neurofilament light (NfL) and glial fibrillary acidic protein (GFAP)).**

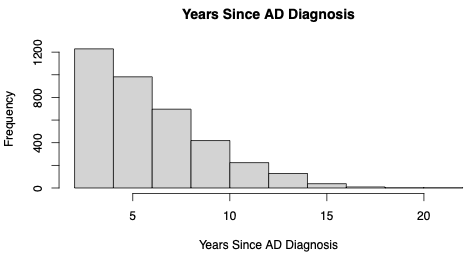

Supplementary Figure 6 – The number of years since AD diagnosis

**Supplementary Methods**

Area under the Curve

To calculate AUC for time-to-event data, it is necessary to set a timepoint at which individuals are classified as cases and controls, we use age 70 (and investigate whether AUC differs using ages 60 and 80).

Microglia gene-set definition

The microglia gene-set for the analysis with biomarkers is defined as follows:

Gosselin et al. defined microglia regions based on GWAS signatures and epigenetic/gene regulatory data. Tansey et al. have redefined this list of SNPs to include established regulatory regions of the genes. SNPs which were within these microglia regions were extracted, and the polygenic risk score was restricted to these specific SNPs to create microglia specific PRS.

Gosselin D, Skola D, Coufal NG, Holtman IR, Schlachetzki JCM, Sajti E, et al. An environment-dependent transcriptional network specifies human microglia identity. Science. 2017;356(6344).

Tansey KE, Cameron D, Hill MJ. Genetic risk for Alzheimer's disease is concentrated in specific macrophage and microglial transcriptional networks. Genome Med. 2018;10(1):14.
